# Development and Validation of Machine Learning Models for Adverse Events after Cardiac Surgery

**DOI:** 10.1101/2025.02.24.25322811

**Authors:** Qingchu Jin, Saeed Amal, Jaime B. Rabb, Felistas Mazhude, Venkatesh Shivandi, Robert S. Kramer, Douglas B. Sawyer, Raimond L. Winslow

**Affiliations:** Roux Institute at Northeastern University, Portland ME, USA; Maine Medical Center, Portland, ME, USA; Koury College of Computer Science, Northeastern University, Boston MA, USA; MaineHealth Institute for Research, Portland ME, USA; Department of Bioengineering, Northeastern University, Boston MA, USA; School of Clinical and Rehabilitation Sciences, Northeastern University, Boston MA, USA

## Abstract

**Importance:** Early recognition of adverse events after cardiac surgery is vital for treatment. However, the widely used Society of Thoracic Surgery (STS) risk model has modest performance in predicting adverse events and only applies <80% of cardiac surgeries.

**Objective:** To develop and validate machine learning (ML) models for predicting outcomes after cardiac surgery.

**Design, setting, and participants:** ML models, referred as Roux-MMC model, were developed and validated using a retrospective cohort extracted from the STS Adult Cardiac Surgery Database (ACSD) at Maine Medical Center (MMC) between January 2012 to December 2021. It was further validated on a prospective cohort of MMC between January 2022 to February 2024. The performance of Roux-MMC model is compared with the STS model.

**Exposure** cardiac surgery

**Main outcomes and measures:** Postoperative outcomes: mortality, stroke, renal failure, reoperation, prolonged ventilation, major morbidity or mortality, prolonged length of stay (PLOS) and short length of stay (SLOS). Primary measure: area under the receiver-operating curve (AUROC).

**Results:** A retrospective cohort of 9,841 patients (median [IQR] age, 67 [59-74] years; 7,127 [72%] males) and a prospective cohort of 2,305 patients (median [IQR] age, 67 [59-73] years; 1,707 [74%] males) were included. In the prospective cohort, the Roux-MMC model achieves performance for prolonged ventilation (AUROC 0.911 [95% CI, 0.887-0.935]), PLOS (AUROC 0.875 [95% CI, 0.848-0.898]), renal failure (AUROC 0.878 [95% CI, 0.829-0.921]), mortality (AUROC 0.882 [95% CI, 0.837-0.920]), reoperation (AUROC 0.824 [95% CI, 0.787-0.860]), SLOS (AUROC 0.818 [95% CI, 0.801-0.835]) and major morbidity or mortality (AUROC 0.859 [95% CI, 0.832-0.884]). The Roux-MMC model outperforms the STS model for all 8 outcomes, achieving 0.020-0.167 greater AUROC. The Roux-MMC model covers all cardiac surgery patients, while the STS model applies to only 65% in the retrospective and 77% in the prospective cohorts.

**Conclusion and relevance:** We developed ML models to predict 8 postoperative outcomes on all cardiac surgery patients using preoperative and intraoperative variables. The Roux-MMC model outperforms the STS model in the prospective cohort. The Roux-MMC model is built on STS ACSD, a data system used in ∼1000 US hospitals, thus, it has the potential to easily applied in other hospitals.

**Key Points:** **Question**. Can a predictive model be developed using preoperative and intraoperative variables from the Society of Thoracic Surgeons (STS) Adult Cardiac Surgery Database (ACSD) to accurately predict adverse events after cardiac surgery?

**Findings.** Machine learning (ML) models developed and validated on a retrospective cohort of 9,841 patients. In the prospective validation cohort, models demonstrated good discrimination for postoperative adverse events including mortality (AUC, 0.882), prolonged ventilation (AUC, 0.911), renal failure (AUC, 0.878), major morbidity or mortality (AUC, 0.859) and prolonged length of stay (AUC, 0.875). ML Models outperform the widely used STS model.

**Meaning**. Predictive models based on preoperative and intraoperative variables from STS ACSD, a data collection system implemented in ∼1000 US hospitals, were developed and validated to predict adverse events after cardiac surgery, allowing for straightforward testing at other institutions.

## Introduction

123.2 per 100,000 people globally and 271.5 per 100,000 people in the US undergo cardiac procedures annually^1^. Postoperative cardiac surgery complications remain a concern, with patients exhibiting the following prevalences for adverse events: 2.9% operative mortality; 10.9% prolonged ventilation; 1.5% stroke; 2.7% renal failure; and 17.4% composite morbidity or mortality^2^. Studies have shown that patient mortality increases significantly if postoperative adverse events occur. This mortality is referred to as failure-to-rescue (FTR). FTR is known to be greater than 30% if more than 2 postoperative adverse events occur^3^. In principle, reliable early prediction of postoperative adverse events before they occur could provide additional intervention time for clinicians to treat and prevent these events to happen, eventually reduce FTR. For example, the rapid response team (RRT), which is the medical emergency care team that intervenes once seeing early signs and symptoms of deterioration, contributed to saving 66,000 lives in the 100,000 Lives Saved Campaign^4^. Early recognition of physiologic deterioration is a vital key for the successful intervention of RRT^5,6^.

The Society of Thoracic Surgeons (STS) risk model is one of the most widely used prediction models for forecasting postoperative outcomes of cardiac surgery patients^2,7^. STS risk model is developed and validated based on ∼670k patients undergo 3 cardiac procedures: coronary artery bypass graft (CABG), valve, and valve+CABG, from the national STS Adult Cardiac Surgery Database (ACSD). The STS risk model has demonstrated an area under the receiver-operating characteristic curve (AUROC) ranging from 0.588 to 0.826 for postoperative outcomes^2^. The modest discriminative performance may contribute to a slow speed in clinical application.

### Justification for a new risk model

We hypothesize that the STS risk model can be improved in 4 perspectives: 1) the STS risk model uses a logistic regression model to generate a risk score. Multiple studies have shown that machine learning (ML) algorithms such as tree-based ensemble methods have better performance than do logistic regression based models^8–10^; 2) the STS risk model was developed using data from patients having undergone CABG, valve, and valve+CABG procedures. 23% of remaining procedures are not covered^11^; 3) the STS risk model was developed using 65 expert-selected preoperative features. The STS ACSD collects hundreds of preoperative variables in addition to these expert-selected preoperative features. Use of these additional features could potentially improve predictive performance. A recent study has shown that use of intraoperative data, which are also available in the STS ACSD, can improve prediction performance in CABG patients^12^; 4) publications on the STS risk model do not report important metrics including sensitivity, specificity, positive predictive value, receiver-operating characteristic curve in the description. Additionally, the parameters of the STS risk model are not publicly available^2^.

In this study, we aim to develop a ML-based prediction model to predict 8 postoperative outcomes for any cardiac surgery patients using preoperative and intraoperative variables that exist in the STS ACSD. We present model performance with metrics including AUROC, sensitivity, specificity and positive predictive value (PPV). We apply SHAP value analysis to interpret the ML model. We show that this model achieves improved performance relative to the STS risk model of 0.020-0.167 AUROC for all outcomes in the prospective validation cohort. Additionally, we showed that 1) ML model has a better performance than logistic regression in all 8 outcomes, 2) intraoperative data has an additive predictive power and 3) using the preoperative variables alone also achieves a better performance than the STS risk model for all 8 outcomes.

## Methods

### Study Design

ML modeling proceeded in three steps: 1) develop a ML model using a data from a retrospective cohort; 2) validate the performance of the ML model using a retrospective patient cohort; and 3) test performance of the ML model on a prospective patient cohort. The Maine Medical Center Institutional Review Board (IRB) and Northeastern University IRB approved this study. This study followed guidelines of the Transparent Reporting of a Multivariable Prediction Model for Individual Prognosis Or Diagnosis + AI (TRIPOD+AI)^13^.

### Data Collection

A retrospective cohort was extracted from the Maine Medical Center STS Adult Cardiac Surgery Database (ACSD)^14^ for patients having cardiac surgeries from January 1, 2012 to December 31, 2021 (9,841 patients). A prospective Maine Medical Center STS ACSD cohort was collected from patients having cardiac surgeries from January 1, 2022 to February. 1, 2024 (2,305 patients). All private health information (PHI) of two cohorts was removed.

### Data Preprocessing and Outcomes

The STS ASCD data collection form has gone through 4 versions from 2012 to 2024. We used variables that exist in the latest version v4.20.2 for these analyses. Data have harmonized across different versions. We systematically screened for potential predictors, excluding (1) variables that are postoperative, (2) variables that fewer than 100 samples are not missing, and (3) variables resulting in one unique value. 373 STS variables (198 preoperative, 170 intraoperative and 5 mixed variables) are selected. Mixed variables contains both preoperative and intraoperative information. All variables are listed in eTable 1 in Supplement 2.

Missing values of continuous variables were imputed using the population mean. Missing values of categorical variables were imputed as a unique category labeled “missing”, recognizing that the absence of data may carry specific significance. Categorical variables were then converted into binary variables by one-hot encoding, leading to 1,451 individual features (Figure 1). 8 postoperative outcomes were selected to align with those predicted using the existing STS risk score model^2,7^: (1) mortality including in-hospital mortality and 30-day discharge mortality; (2) renal failure; (3) stroke; (4) prolonged ventilation; patients having postoperative pulmonary ventilation >24 hrs (prolonged ventilation); (5) reoperation for bleeding, tamponade, or any cardiac reason; (6) major morbidity or mortality - an ensemble outcome in which any outcome of (1) to (5) occur; (7) prolonged length of stay (PLOS, LOS>14 days); and (8) short length of stay (SLOS, LOS <6 days).

**Figure 1.**
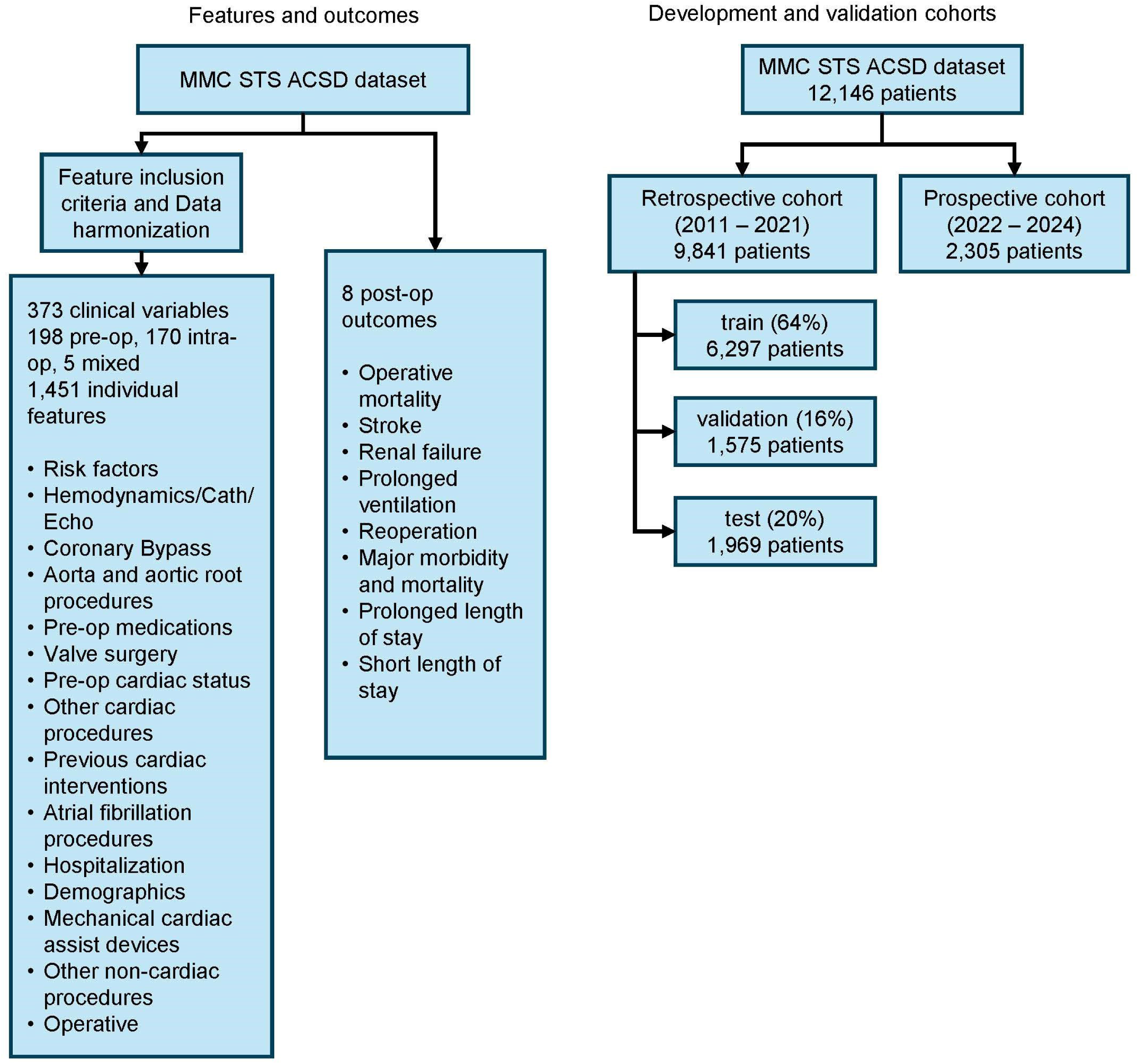
Feature selection and cohort development. The left diagram shows the step for selecting 373 STS clinical variables (198 preoperative, 170 intraoperative and 5 mixed) as predictors and 8 postoperative outcomes. The right diagram shows the split of the cohorts.

### Model Development

Figure 1 shows that the retrospective cohort was randomly sampled into the training set (64%, 6,297 patients), validation set (16%, 1,575 patients) and test set (20%, 1,969 patients). The model was trained on the training set. The validation set was used to optimize hyperparameters of the ML models. We used the Random Forest algorithm^15^, an ensemble tree-based algorithm, to develop ML models. For each outcome, we develop a Random Forest model for prediction. 4 hyperparameters with the ranges are used for searching:

1. number of estimators: [50, 1000]
2. max depth of tree: [None, 10, 20, 30]
3. min samples required to split a node: [2, 11]
4. min samples required at a leaf node: [1, 5].

We randomly generated 30 hyperparameter sets and fed into the model. The model performance is evaluated by the area under the operating receiver curve (AUROC) on the validation set. The hyperparameter set yielding the highest AURPC was selected, and the final model was retrained on a combined dataset of the training and validation sets using the optimal hyperparameter set. The predicted adverse event risk will be represented as continuous value between 0 and 1, we set a threshold on the risk to make binary prediction.

Model performance was evaluated on the test set and the prospective cohort using AUROC, sensitivity, specificity, positive predictive value (PPV), negative predictive value (NPV), accuracy and area under precision-recall curve (AUPRC) as metrics. Metrics other than AUROC and AUPRC rely on the selection of a specific threshold on the risk score for predicting an adverse event. We chose two thresholds for evaluation: (1) the threshold that maximizes the Yonden’s J statistics (= sensitivity + specificity −1), which balances sensitivity and specificity, and (2) the threshold that maximizes the F1 score (harmonic mean of PPV, namely precision, and sensitivity, namely recall), balancing sensitivity and PPV.

### Other Machine Learning Models and STS Risk Model

Besides Random Forest, we also examined CatBoost^16^ and XGBoost^17^, two widely-used ML algorithms for predicting postoperative outcomes. We also, developed a logistic regression-based classifier (note that the parameters of the STS ACSD classifier are not published). The STS ACSD data collection system automatically calculates the predicted risk for those 8 outcomes using the previously developed STS risk model^2,7^. All patients undergone CABG, valve and CABG+valve in cohorts were used to calculate the AUROC of the STS risk model.

### Model only using Preoperative Variables

We examined the predictive power of preoperative variables by developing Random Forest models based only on use of these preoperative variables. We evaluate model performance on the retrospective test set and the prospective cohort. The added predictive power of intraoperative variables is quantified by the difference in AUROC between models only using preoperative variables and those using both preoperative and intraoperative variables.

### Model Explanation

To improve the interpretation of the predicted risk, Shapley additive explanation (SHAP) value were used^18^. SHAP is a model-agnostic explanation method that can be used to understand the importance of specific features for model prediction. A positive SHAP value for a feature indicates that the feature significantly contributes to increase the predicted risk, and vice versa. To calculate the feature importance of a categorical variable, the absolute sum of the SHAP values of all its one-hot encoded features is taken to determine the contribution.

### Statistical Test

Delong test^19^ is performed to compare AUROCs across two models, with a significance threshold set at 0.05.

## Results

A retrospective cohort of 9,841 patients (7,127 males, 72%) with a median age 67 years: (IQR: 59-74) from January 2012 to February 2021 and a prospective cohort of 2,305 patients (1,707 males, 74%), with a median age 67 years (IQR: 59-73) from January 2022 to February 2024 are used to develop and validate the model (Table 1).

**Table 1.** Selected STS variables and outcome variables for retrospective and prospective cohorts.

|  | Retrospective cohort<br>Jan 2012 – Dec 2021<br>N = 9,841 | Prospective Cohort<br>Jan 2022 – Feb 2024<br>N = 2,305 |
| --- | --- | --- |
| Variables |  |  |
| Age | 67 (59, 74) | 67 (59, 73) |
| Male | 7,127 (72%) | 1,707 (74%) |
| Body mass index | 28.9 (25.4, 32.7) | 28.6 (25.3, 32.7) |
| Preop ejection fraction | 56 (50, 62) | 58 (50, 62) |
| Clinical Status in OR (%) |  |  |
| Elective | 4,169 (42%) | 1,004 (44%) |
| Emergent | 453 (4.6%) | 58 (2.5%) |
| Emergent Salvage | 10 (0.1%) | 4 (0.2%) |
| Urgent | 5,203 (53%) | 1,238 (54%) |
| Types of procedures |  |  |
| Isolated CABG | 3,836 (39%) | 1,172 (51%) |
| Isolated Valve | 1,501 (15%) | 373 (16%) |
| Valve + CABG | 1,089 (11%) | 242 (10%) |
| Other | 3,415 (35%) | 518 (23%) |
| Outcomes |  |  |
| Mortality | 342 (3.5%) | 74 (3.2%) |
| Stroke | 198 (2.0%) | 37 (1.6%) |
| Renal failure | 190 (1.9%) | 55 (2.4%) |
| Prolonged ventilation | 1,000 (10%) | 185 (8.0%) |
| Reoperation | 547 (5.6%) | 133 (5.8%) |
| Major morbidity or mortality | 1,536 (16%) | 290 (13%) |
| Prolonged LOS | 777 (7.9%) | 193 (8.4%) |
| Short LOS | 4,134 (42%) | 1,140 (49%) |

Continuous variable: median (IQR), categorical variables: n (%), 1 patient in retrospective cohort misses
length of stay. LOS: length of stay

The Random Forest model was developed with 373 preoperative and intraoperative STS variables on the training and validation set in the retrospective cohort. This model is referred to as the Roux-MMC model. Figures 2A-C show that the Roux-MMC model achieves 0.858 [95%CI 0.812-0.902] AUROC for mortality, 0.834 [95%CI 0.806-0.861] AUROC for major morbidity or mortality and 0.899 [95%CI 0.875-0.922] AUROC for prolonged ventilation, all significantly better than STS risk model. Figure 2D shows that except the PLOS, the Roux-MMC model has 0.009-0.096 increase of AUROC than the STS risk score model. In the prospective cohort validation, the Roux-MMC model achieves 0.882 [95%CI 0.837-0.920] AUROC for mortality, 0.859 [95%CI 0.832-0.884] AUROC for major morbidity or mortality and 0.911 [95%CI 0.887-0.935] AUROC for prolonged ventilation, also significantly better than the STS risk model (Figures 2E-G). Figure 2H shows that the Roux-MMC model has a better performance than STS risk model on all 8 outcomes with an 0.020-0.167 increase of AUROC. This demonstrates the strong predictive power of Roux-MMC model particularly on predicting mortality, major morbidity or mortality and prolonged ventilation. eTables 2-5 in Supplement 1 demonstrate performance metrics including AUROC, sensitivity, specificity, PPV and NPV for each outcome for two datasets. eTable 6 in Supplement 1 reports the AUROC of STS risk model on retrospective and prospective cohorts.

**Figure 2.**
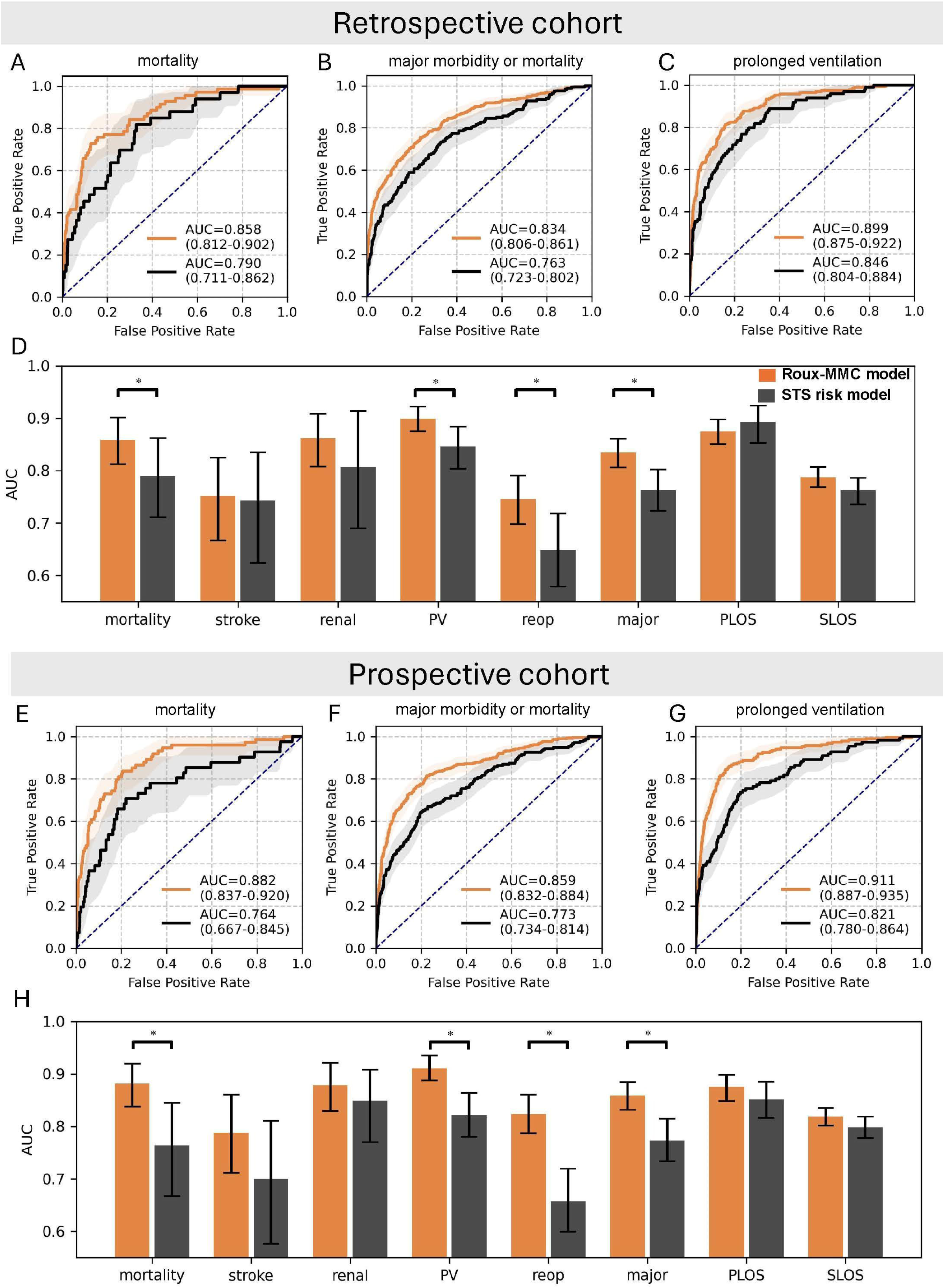
Model performance on the test set of retrospective cohort and prospective cohort. For the retrospective cohort, the receiver operating characteristics (ROC) curve is presented for Roux-MMC model (orange) and STS risk model (black) for (A) mortality, (B) major morbidity or mortality and (C) prolonged ventilation. The ribbon on the ROC curve is the 95% CI. (D). AUROC for 8 outcomes between Roux-MMC model and STS risk model. (E-H) shows the similar ROC curves and AUROCs as (A-D) on the prospective cohort. PV: prolonged ventilation. PLOS: prolonged length of stay, SLOS: short length of stay. Delong test was performed to compare AUROCs across two models for each outcome. Significance threshold is 0.05.

### Added value of intraoperative variables

We then developed a Random Forest model only based on preoperative variables (198 preoperative variables and 5 mixed variables, where intraoperative information has been removed for 5 mixed variables). Table 2 compares the AUROC between Random Forest with only preoperative data and the Roux-MMC model (with both preoperative and intraoperative variables). Roux-MMC model has a 0.009-0.034 greater AUROC than the Random Forest with pre-op variables for renal failure, prolonged ventilation, reoperation, major morbidity or mortality, PLOS and SLOS in retrospective cohort, and has a 0.009-0.027 greater AUROC in mortality, stroke, renal failure, prolonged ventilation, reoperation, major morbidity or mortality and SLOS. It indicates that intra-op has a marginal added predictive value for most outcomes. Additionally, Random Forest only with pre-op variables achieved 0.015-0.068 greater AUROC than STS risk score for all outcomes except PLOS in the retrospective cohort and 0.008-0.140 greater AUROCs for all outcomes in the prospective cohort. STS risk model also builds a risk score on the preoperative variables (65 expert selected preoperative variables). Our preoperative model outperforms the STS risk model, indicating that other preoperative variables have an added value.

**Table 2.** AUROC comparison between different STS variable sets on both retrospective and prospective cohorts: pre-op only and pre-op + intra-op.

|  | Retrospective cohort |  |  | Prospective cohort |  |  |
| --- | --- | --- | --- | --- | --- | --- |
|  | RF<br>Pre-op | Roux-MMC<br>(Pre-op +<br>Intra-op) | STS risk<br>model | RF<br>Pre-op | Roux-MMC<br>(Pre-op +<br>Intra-op) | STS risk<br>model |
| mortality | 0.858 | 0.858 | 0.790 | 0.861 | 0.882 | 0.764 |
| stroke | 0.764 | 0.752 | 0.743 | 0.767 | 0.788 | 0.700 |
| renal | 0.853 | 0.862 | 0.807 | 0.850 | 0.878 | 0.849 |
| PV | 0.886 | 0.899 | 0.846 | 0.902 | 0.911 | 0.821 |
| reoperation | 0.711 | 0.745 | 0.649 | 0.797 | 0.824 | 0.657 |
| major | 0.821 | 0.834 | 0.763 | 0.849 | 0.859 | 0.773 |
| PLOS | 0.848 | 0.875 | 0.893 | 0.879 | 0.875 | 0.851 |
| SLOS | 0.777 | 0.787 | 0.762 | 0.806 | 0.818 | 0.798 |

RF: Random Forest; PV: prolonged ventilation; renal: renal failure; major: major morbidity or mortality; PLOS: prolonged length of stay; SLOS: short length of stay

### Model Interpretability

The importance of each feature importance for predicting major morbidity or mortality was analyzed with SHAP values based on the prospective test cohort. Figure 3A shows the top 20 most important features ranked by the sum of the absolute value of SHAP values across all patients in the prospective cohort. The top 20 most important features for predicting prolonged ventilation and mortality are shown in eFigures 1-2 in Supplement 1. Figure 3B shows how the variable values of the top 8 predictive features are associated with the SHAP value. Missing values of IABP (intra-aortic balloon pump), IABP timing and ECMO (extracorporeal membrane oxygenation) indicate that those procedures were not performed. The greater SHAP value indicates a strong contribution to increasing the predicted risk for the variable.

**Figure 3.**
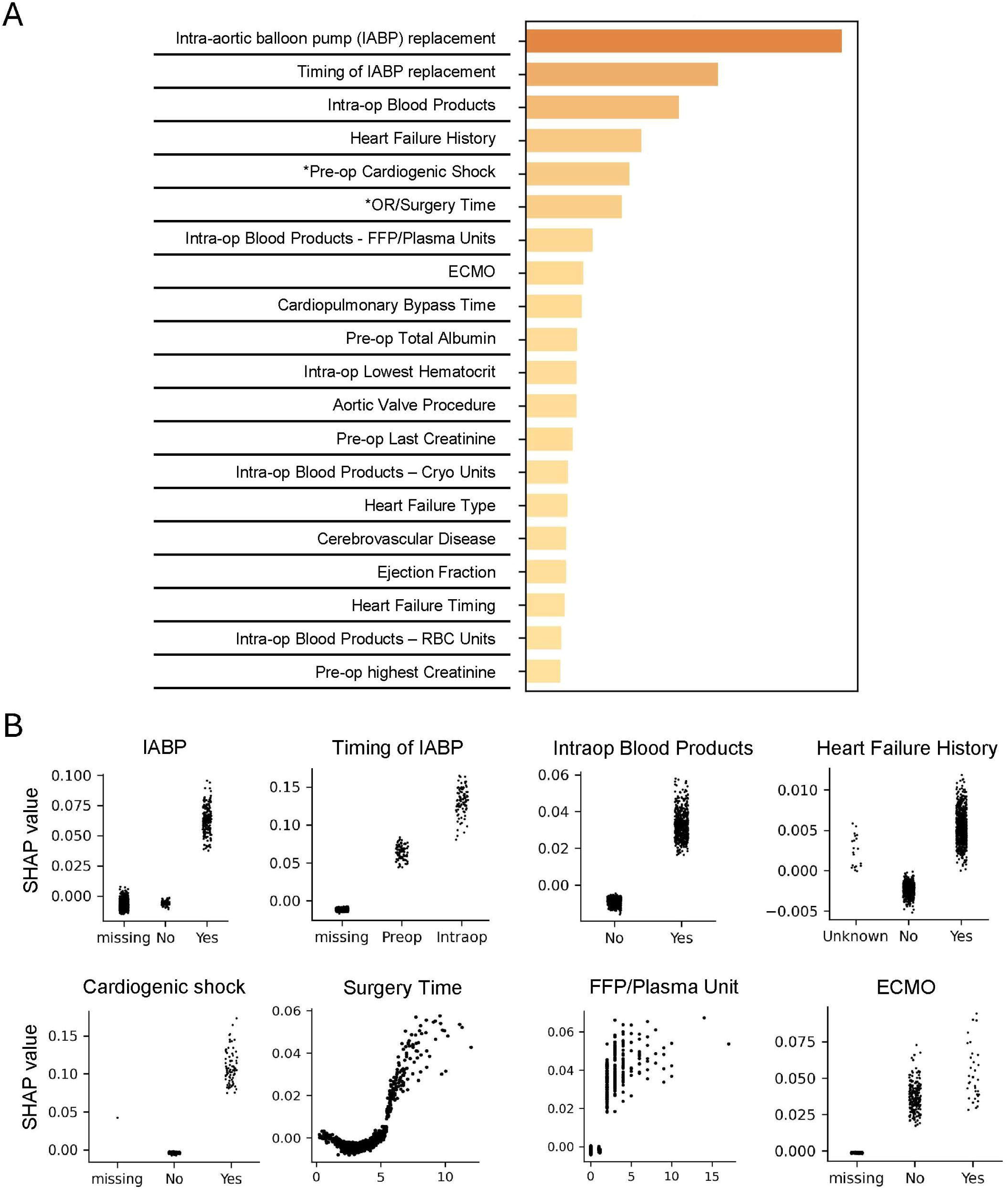
Important variables for major morbidity or mortality prediction based on SHAP value. A. The top 20 most important variables ranked by SHAP value for predicting major morbidity or mortality. B. For the top 8 predictive variables, the SHAP value is shown for each patient (y axis) with the variable value (x axis). IABP: intra-aortic balloon pump; FFP: Fresh Frozen Plasma. ECMO: extracorporeal membrane oxygenation.

## Discussion

In this prognostic study, we developed the Random Forest-based Roux-MMC model to predict 8 postoperative adverse events for all cardiac surgery patients with STS ACSD preoperative and intraoperative variables. We showed that, in the validation of the prospective cohort (2022-2024), we achieved 0.020-0.167 greater AUROCs of all 8 outcomes than does the STS risk model, one of the most widely used operative risk calculators for cardiac surgery patients^20^. Particularly, Roux-MMC model achieves 0.911 AUROC for prolonged ventilation, 0.882 for mortality, 0.878 for renal failure, 0.875 for PLOS and 0.859 for major morbidity or mortality, all greater than STS risk model.

Roux-MMC model has 3 primary improvements compared with STS risk model (eTable 7 in Supplement 1). (1) Roux-MMC model used the Random Forest, a tree-based ensemble ML algorithm to develop the model. This enables the algorithm to capture nonlinear relationships between features and outcomes. We also explored whether use of different ML algorithms leads to improved prediction performance on the same 373 STS variables. CatBoost^16^, XGBoost^17^ and logistic regression models were developed for each outcome. eTable 8 in Supplement 1 shows that, for each outcome, Random Forest, CatBoost and XGBoost all achieved similar performance, except for stroke prediction, where CatBoost has 0.05 lower AUROC than does XGBoost and Random Forest. The logistic regression model has a consistent 0.05-0.08 lower AUROC for all 8 outcomes relative to all other ML models. These results show that tree-based ML models can capture complex association between predictors and outcomes, thus outperform the logistic regression. (2) STS risk model does not apply to all procedures. In our dataset, more than one fifth of cardiac surgeries are not covered (23% for prospective cohort, 35% for retrospective cohort). In contrast, Roux-MMC model applied to all cardiac surgeries. eTable 9 in Supplemen 1 shows the Roux-MMC model AUROC for three major procedures: CABG, valve and CABG+valve and the other surgery procedures. Roux-MMC model achieves good AUROC (0.913 for prolonged ventilation, 0.878 for mortality, 0.859 for SLOS and 0.856 for major morbidity or mortality) in the other surgery procedure - surgeries without established risk scores. Except SLOS, the Roux-MMC model achieves better performance than STS risk model on all 7 outcomes for patients undergone 3 major cardiac procedures. (3) STS risk model focused on an expert-selected 65 preoperative variables. Roux-MMC model uses 198 preoperative, 170 intraoperative and 5 mixed variables. Table 2 shows the added value of intraoperative data, consistent with other studies^12,21^. Additionally, Table 2 shows that in the prospective cohort, only using preoperative variables also achieves better performance than STS risk model across all 8 outcomes. This indicates that other preoperative variables besides the 65 expert-selected variables can add values for prediction. Besides the model difference, STS risk model does not report important model performance metrics such as sensitivity, specificity and positive predictive value. eTables 2-5 in Supplement 1 shows the sensitivity, specificity, PPV, NPV, accuracy and AUPRC for each outcome on retrospective and prospective cohorts.

There exist some efforts to improve some of three aspects (eTable 7 in Supplement 1) of the STS risk model. Kilic et al. developed a XGBoost model to predict mortality on three cardiac procedures: CABG, valve and valve+CABG with preoperative variables^9^. They showed a modest improvement of XGBoost compared with STS risk model. Mori et al. developed a XGBoost model to predict 7 outcomes only on CABG with preoperative and intraoperative variables^12^. They demonstrated that intraoperative variables improved the prediction on all 7 outcomes. Recently, Ong et al. develop ML-driven models for predicting operative mortality specifically on cardiac surgical procedures that STS risk model doesn’t cover with preoperative variables. Our work is the first study, to our knowledge, to address all three aspects of surgical risk prediction for cardiac surgery, by developing a ML model on all cardiac surgeries with both preoperative and intraoperative variables. Additionally, it predicts 8 outcomes. Many studies only focused on predicting mortality^11,22^. Other postoperative outcomes such as renal failure and stroke are also clinically important to predict. Studies have shown that patient mortality increases significantly if postoperative adverse events occur^3^.

We applied SHAP value to interpret the model, obtaining the top 20 features contributing to the prediction. Figure 3B shows the correlation between the variable value and SHAP value of top 8 predictive features for predicting the major morbidity or mortality. These 8 features and the correlation with the contribution are consistent with the literatures: timing of IABP replacement^23^, intraoperative blood transfusion^24^, heart failure history^25^, cardiogenic shock^26^, surgery time^27^ and ECMO^28^.

An additional advantage of the Roux-MMC model is its development based on the variables from STS ACSD, one of the widely used data collection system available in >1000 US hospital and medical institutions, representing >90% US adult cardiac surgery hospitals^29^. Thus, the model can be tested on numerous hospitals with minimal additional data collection. This enables the Roux-MMC model a powerful predictive tool not only in Maine Medical Center but also for a wide range of US hospitals.

### Limitations

The Roux-MMC model has been validated in both retrospective and prospective cohorts in a single institution. The limited sample size let the improvement of AUROC for renal failure, stroke and length of stay prediction not significant (Figures 2D and 2H). In future, we plan to externally validate our model with other institutions to examine the model generalizability and increase the power of the analysis. Roux-MMC model is developed based on the STS ACSD. Additional variables in the electronic healthcare record (EHR) data may further improve the prediction. Recently, Weiss et al. develop a XGBoost model to predict mortality for cardiac surgery patients with EHR, with 0.978 AUROC. This demonstrates the strong potential of EHR data^22^.

## Conclusions

In this prognostic study, we developed a ML driven risk score, Roux-MMC model, accurately predicting 8 postoperative outcomes of all cardiac surgery patients. The model is predicated on 373 preoperative and intraoperative STS variables. The Roux-MMC model demonstrates a superior predictive performance than STS risk model on all 8 outcomes. We also showed that intraoperative variables have added value in predicting postoperative outcomes. Further research on external validation in other medical institutions is needed to prove the model generalizability.

## Supporting information

Supplement eTable 1

## Data Availability

Upon request to Drs. Raimond L. Winslow and Qingchu Jin with a scientific question, de-identified data may be share

## Acknowledgement

We would like to thank for the funding support of the HEART Impact Engine of the Northeastern University. Drs. Robert Kramer and Douglas Sawyer receive funding from NIH/NIGMS UM1Hl147371, Dr. Douglas Sawyer is supported by NIH/NIGMS P20GM139745.

