## Supplemental materials for "Development and Validation of Machine Learning Models for Adverse Events after Cardiac Surgery"

**eTable 1.** Description of 373 preoperative and intraoperative STS ACSD variables (in the separated supplement excel file)

**eFigure 1.** Top 20 most important variables for predicting mortality

**eFigure 2.** Top 20 most important variables for predicting prolonged ventilation

**eTable 2.** Roux-MMC model performance metrics on the retrospective cohort. The threshold is chosen to maximize the Yonden’s J statistics

**eTable 3.** Roux-MMC model performance metrics on the retrospective cohort. The threshold is chosen to maximize the F1 score

**eTable 4.** Roux-MMC model performance metrics on the prospective cohort. The threshold is chosen to maximize the Yonden’s score

**eTable 5.** Roux-MMC model performance metrics on the prospective cohort. The threshold is chosen to maximize the F1 score

**eTable 6.** AUROC of STS risk score model performance (only on patients undergone CABG, valve and CABG+valve)

**eTable 7.** The major improvement of Roux-MMC model comparing with STS risk model

**eTable 8.** AUROC of the test set of the retrospective cohort across 4 different machine learning algorithms

**eTable 9.** AUROC of Roux-MMC model for different type of surgeries on the prospective cohort

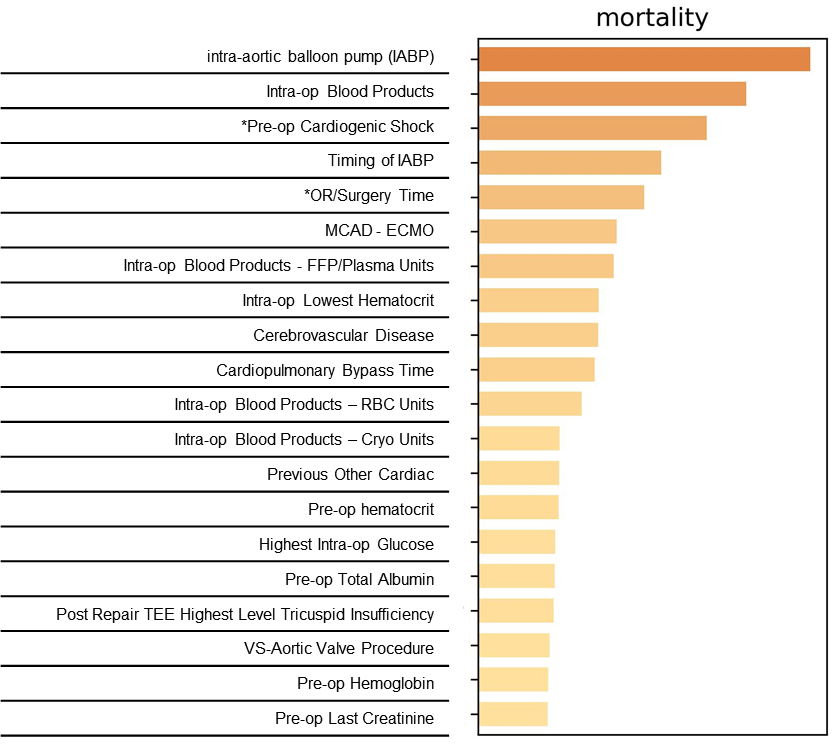

**eFigure 1. Top 20 most important variables ranked by SHAP value for predicting mortality**

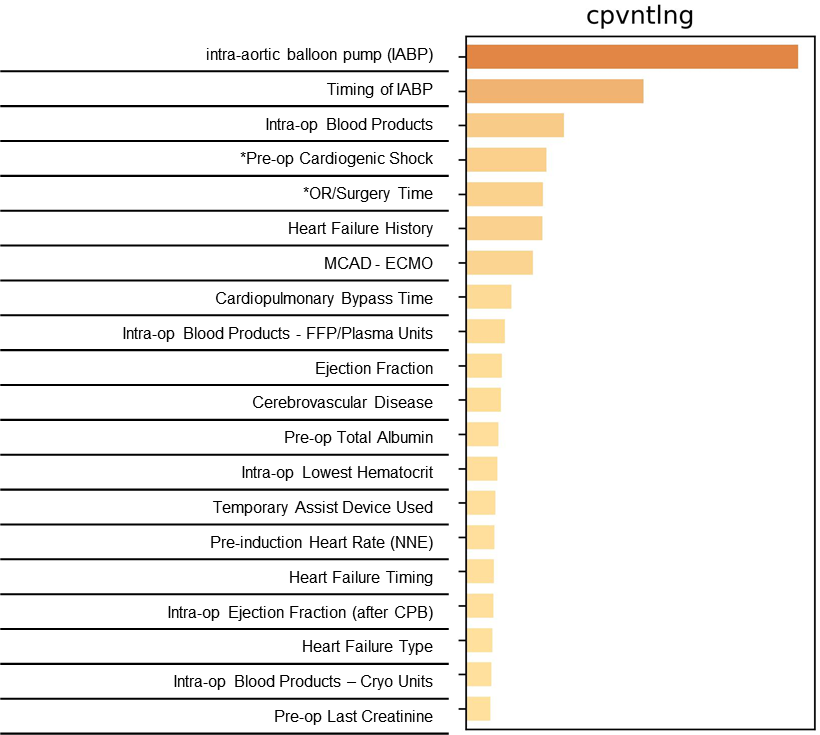

**eFigure 2. Top 20 most important variables ranked by SHAP value for predicting prolonged ventilation**

**eTable 2. Roux-MMC model performance metrics on the retrospective cohort. The threshold is chosen to maximize the Yonden’s J statistics.**

|  | mortality | stroke | renal | PV | reop | major | PLOS | SLOS |
| --- | --- | --- | --- | --- | --- | --- | --- | --- |
| AUROC | 0.858 | 0.752 | 0.862 | 0.899 | 0.745 | 0.834 | 0.875 | 0.787 |
| sensitivity | 0.729 | 0.692 | 0.730 | 0.814 | 0.624 | 0.728 | 0.787 | 0.771 |
| specificity | 0.857 | 0.734 | 0.812 | 0.834 | 0.740 | 0.786 | 0.803 | 0.678 |
| PPV | 0.158 | 0.050 | 0.069 | 0.349 | 0.132 | 0.384 | 0.280 | 0.641 |
| NPV | 0.988 | 0.992 | 0.994 | 0.976 | 0.969 | 0.940 | 0.975 | 0.798 |
| threshold | 0.060 | 0.020 | 0.030 | 0.120 | 0.060 | 0.150 | 0.100 | 0.420 |
| AUPRC | 0.283 | 0.080 | 0.109 | 0.572 | 0.207 | 0.603 | 0.468 | 0.706 |
| accuracy | 0.853 | 0.733 | 0.811 | 0.832 | 0.733 | 0.777 | 0.802 | 0.718 |

PPV: positive predictive value; NPV: negative predictive value; AURPC: area under precision-recall curve

**eTable 3. Roux-MMC model performance metrics on the retrospective cohort. The threshold is chosen to maximize the F1 score.**

|  | mortality | stroke | renal | PV | reop | major | PLOS | SLOS |
| --- | --- | --- | --- | --- | --- | --- | --- | --- |
| sensitivity | 0.386 | 0.128 | 0.270 | 0.588 | 0.316 | 0.502 | 0.529 | 0.771 |
| specificity | 0.977 | 0.987 | 0.974 | 0.959 | 0.935 | 0.945 | 0.931 | 0.678 |
| PPV | 0.380 | 0.167 | 0.164 | 0.613 | 0.234 | 0.627 | 0.426 | 0.641 |
| NPV | 0.977 | 0.982 | 0.986 | 0.955 | 0.956 | 0.912 | 0.953 | 0.798 |
| threshold | 0.160 | 0.130 | 0.100 | 0.340 | 0.120 | 0.330 | 0.210 | 0.420 |
| accuracy | 0.956 | 0.970 | 0.960 | 0.923 | 0.898 | 0.877 | 0.895 | 0.718 |

**eTable 4. Roux-MMC model performance metrics on the prospective cohort. The threshold is chosen to maximize the Yonden’s J statistics**

|  | mortality | stroke | renal | PV | reop | major | PLOS | SLOS |
| --- | --- | --- | --- | --- | --- | --- | --- | --- |
| AUROC | 0.882 | 0.788 | 0.878 | 0.911 | 0.824 | 0.859 | 0.875 | 0.818 |
| sensitivity | 0.811 | 0.568 | 0.800 | 0.854 | 0.789 | 0.724 | 0.803 | 0.844 |
| specificity | 0.800 | 0.851 | 0.849 | 0.855 | 0.690 | 0.858 | 0.817 | 0.627 |
| PPV | 0.119 | 0.058 | 0.115 | 0.339 | 0.135 | 0.423 | 0.287 | 0.689 |
| NPV | 0.992 | 0.992 | 0.994 | 0.985 | 0.982 | 0.956 | 0.978 | 0.804 |
| threshold | 0.060 | 0.040 | 0.060 | 0.190 | 0.060 | 0.250 | 0.160 | 0.360 |
| AUPRC | 0.342 | 0.117 | 0.162 | 0.540 | 0.305 | 0.565 | 0.412 | 0.790 |
| accuracy | 0.800 | 0.846 | 0.848 | 0.855 | 0.695 | 0.841 | 0.816 | 0.734 |

**eTable 5. Roux-MMC model performance metrics on the prospective cohort. The threshold is chosen to maximize the F1 score**

|  | mortality | stroke | renal | PV | reop | major | PLOS | SLOS |
| --- | --- | --- | --- | --- | --- | --- | --- | --- |
| sensitivity | 0.311 | 0.189 | 0.455 | 0.595 | 0.489 | 0.634 | 0.622 | 0.902 |
| specificity | 0.990 | 0.993 | 0.958 | 0.961 | 0.926 | 0.920 | 0.909 | 0.567 |
| PPV | 0.511 | 0.304 | 0.208 | 0.570 | 0.288 | 0.532 | 0.383 | 0.671 |
| NPV | 0.977 | 0.987 | 0.986 | 0.964 | 0.967 | 0.946 | 0.963 | 0.855 |
| threshold | 0.250 | 0.140 | 0.120 | 0.380 | 0.140 | 0.320 | 0.250 | 0.320 |
| accuracy | 0.968 | 0.980 | 0.946 | 0.931 | 0.901 | 0.884 | 0.885 | 0.732 |

**eTable 6. AUROC of STS risk score model performance (only on patients undergone CABG, valve and CABG+valve)**

|  | Retrospective cohort | Prospective cohort |
| --- | --- | --- |
| mortality | 0.790 (0.711 – 0.862) | 0.764 (0.667 – 0.845) |
| stroke | 0.743 (0.624 – 0.834) | 0.700 (0.576 – 0.811) |
| renal | 0.807 (0.690 – 0.914) | 0.849 (0.770 – 0.908) |
| PV | 0.846 (0.804 – 0.884) | 0.821 (0.780 – 0.864) |
| reop | 0.649 (0.579 – 0.719) | 0.657 (0.600 – 0.719) |
| major | 0.763 (0.723 – 0.802) | 0.773 (0.734 – 0.814) |
| PLOS | 0.893 (0.853 – 0.923) | 0.851 (0.816 – 0.885) |
| SLOS | 0.762 (0.735 – 0.786) | 0.798 (0.778 – 0.818) |

**eTable 7. The major improvement of Roux-MMC model comparing with STS risk model**

|  | **Roux-MMC model** | **STS risk model** |
| --- | --- | --- |
| algorithm | Random forest | Logistic regression |
| Cardiac procedures | All cardiac procedures | CABG, valve, valve+CABG (<80% of cardiac surgeries) |
| Variable | 373 preoperative and intraoperative variables | 65 preoperative variables |

**eTable 8. AUROC of the test set of the retrospective cohort across 4 different machine learning algorithms**

|  | Random Forest  (Roux-MMC model) | CatBoost | XGBoost | logistic |
| --- | --- | --- | --- | --- |
| mortality | 0.858 | 0.867 | 0.853 | 0.810 |
| stroke | 0.752 | 0.704 | 0.754 | 0.709 |
| renal | 0.862 | 0.858 | 0.873 | 0.792 |
| PV | 0.899 | 0.895 | 0.901 | 0.835 |
| reoperation | 0.745 | 0.745 | 0.729 | 0.653 |
| major | 0.834 | 0.837 | 0.837 | 0.776 |
| PLOS | 0.875 | 0.882 | 0.882 | 0.824 |
| SLOS | 0.787 | 0.791 | 0.794 | 0.730 |

RF: Random Forest; PV: prolonged ventilation; renal: renal failure; major: major morbidity or mortality; PLOS: prolonged length of stay; SLOS: short length of stay

**eTable 9. AUROC of Roux-MMC model for different type of surgeries on the prospective cohort**

|  | CABG, valve or valve+CABG | others |
| --- | --- | --- |
| mortality | 0.886 | 0.878 |
| stroke | 0.714 | 0.823 |
| renal | 0.883 | 0.833 |
| PV | 0.901 | 0.913 |
| reop | 0.827 | 0.779 |
| major | 0.847 | 0.856 |
| PLOS | 0.873 | 0.846 |
| SLOS | 0.800 | 0.859 |
